# A Statistical Analysis Of CoV-19 Positive Test Frequency Data Indicates A Need For Greater Attention To CoV-19 Test Quality And Pre-Wuhan Cov-19 Prevalence

**DOI:** 10.1101/2020.04.24.20078402

**Authors:** James L. Sherley

## Abstract

Increased attention to analysis of SARS-CoV-2 (CoV-19) positive test frequency data is essential for achievement of better knowledge of the natural history of the virus in human populations, improved accuracy of CoV-19 epidemiological data, and development of public response policies that are better crafted to address the current CoV-19-induced global crisis. A statistical analysis of currently available positive test frequency data reveals a surprisingly uniform relationship between the number of CoV-19 test performed and the number of positive tests obtained. The uniformity is particularly striking for United States CoV-19 test data. Such observations warrant closer evaluation of other factors, besides virus spread, that may also contribute to the nature of the coronavirus pandemic. These include indigenous CoV-19 and the quality of CoV-19 testing.

---

Recent reports, especially in the lay press, have focused primarily on the rise in the number of individuals testing positive for CoV-19 (SARS-CoV-2). Less attention has been given to evaluating statistical relationships between the number of positive tests and the number of tests performed. Statistical interrogation of positive test frequency in the current frenzy of test analyses could provide clues to aspects of the current crisis that would better guide response strategies and policy. In particular, such analyses may inform an emerging question regarding the natural history of CoV-19 in human populations. Namely, the possibility that the virus is not new to human populations; but has only now come into awareness and detection because of events occurring recently in Wuhan, China (1). This issue is particularly important for attention in situations like the present, when non-clinical measures of disease incidence dominate epidemiological assessments.

World Health Organization (WHO) and U.S. Centers for Disease Control and Prevention (CDC) data organized and posted online make it possible to perform statistical analyses of CoV-19-positive test frequency data from 45 countries and sub-regions (2; Fig. 1A and 1B) and 45 U.S. states and the District of Columbia (3; Fig. 1C and 1D). Fig. 1 provides frequency plots (Fig. 1A and Fig. 1C) and correlation plots (Fig. 1B and Fig. 1D) of these respective data. For global data, the mean positive test frequency is 8.0% (Fig. 1A; p < 0.0001; 95% CI 5.2% to 11%; median = 6.0%). A significant positive correlation exists between the number of tests conducted and the number of positive tests results obtained (Fig. 1B; p < 0.01). The Pearson correlation coefficient (R^2^) indicates that about 58% of the variance observed in the number of positive tests obtained can be explained by differences in the number of tests performed in different countries. The remaining 42% of the variance is predicted to be due to other factors.

**Figure 1.**
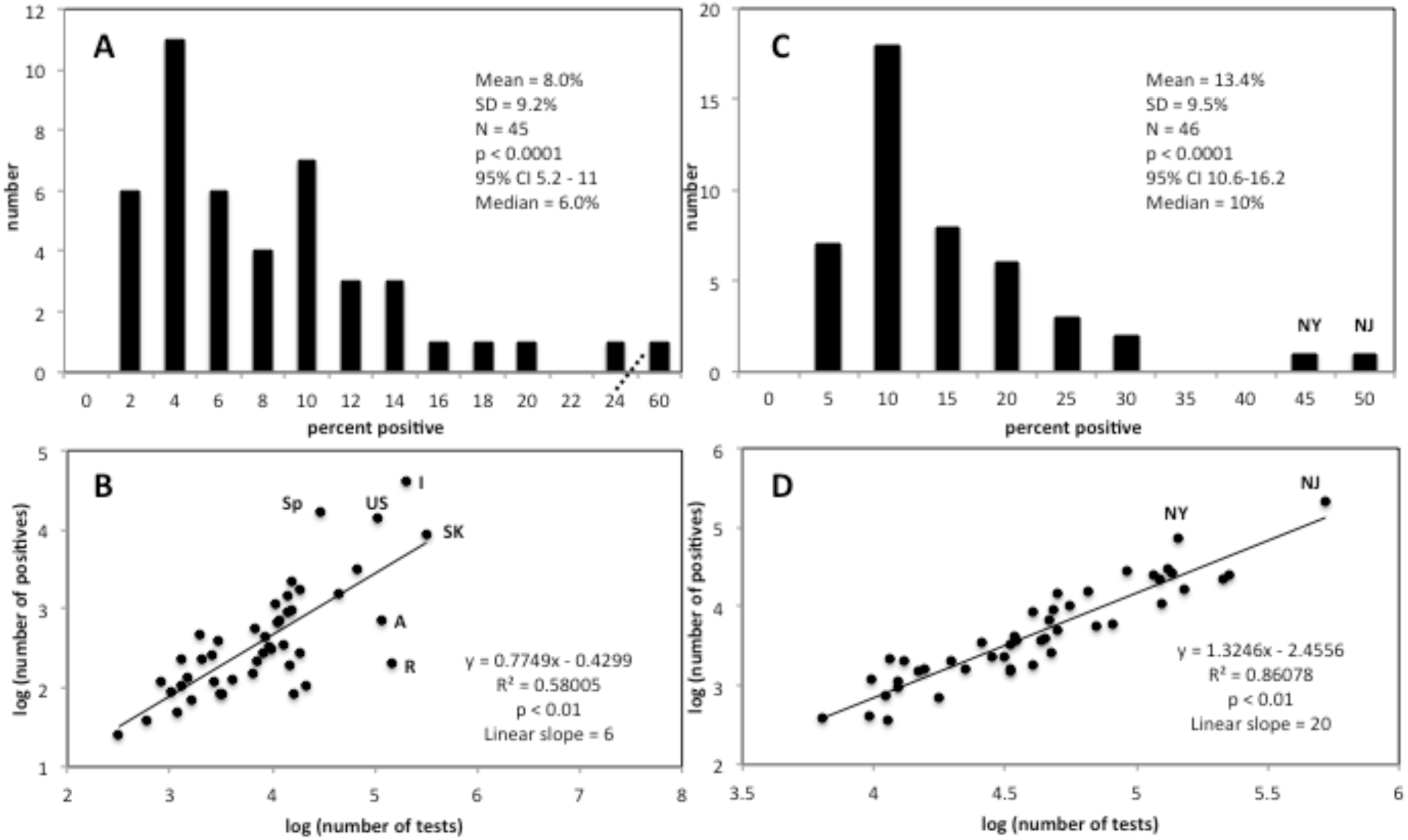
Analysis of global and U.S. Cov-19 positive test frequency data.

The countries that deviate the most from the axis of the data in Fig. 1B include Spain (Sp), Italy (I), and the United States (US), showing the highest positive test frequencies at 56.6%, 20%, and 13.4%, respectively, compared, for instance, to South Korea (SK) at 2.7%. Examples of countries differing with lower positive test frequencies are Australia (A) and Russia (R), with frequencies of 0.14% and 0.61%, respectively. These deviations indicate that other important factors may act in these particular countries, including differences in the rate of CoV-19 spread. However, factors related to testing quality could also be responsible, including test specificity in the case of higher positive test frequencies and test sensitivity in the case of lower positive test frequencies.

The analysis of U.S. states data raises even greater concern (Fig. 1C and Fig 1D) of effects related to testing, but not to CoV-19 spread. The mean positive test frequency is 13.4% (Fig. 1C; p < 0.0001; 95% CI 10.6% to 16.2%; median = 10%), equivalent to the value from WHO data. However, based on the Pearson correlation coefficient, 86% of the variance can be explained by differences in the number of tests performed (Fig. 1D). It seems unlikely that such a high correlation, uniformly across the continental U.S., could be explained by CoV-19 spread from the event in Wuhan. Even the states of New Jersey (NJ) and New York (NY), which have apparently distinctively higher positive test frequencies, can be accounted for by chance occurrence within the log-normal distributions observed for the positive test frequency data (Fig. 1A and Fig. 1C).

At a very high rate, positive CoV-19 tests in the U.S. have no identifiable source of transmission. As of this writing, an estimated 97.6% of positive tests in the U.S. fall into this category (4). Such a high rate of unknown route of presumed transmission may indicate that the high positive test frequency in the U.S. has two other responsible factors. The first is a possible high false positive rate in U.S. testing. Since the instructions disseminated by the U.S. CDC allow a test error rate as a high as 10% (5), this factor is a serious possibility that warrants greater attention. The second possible factor is a significant frequency of CoV-19 in the U.S. population prior to the event in Wuhan, China. Analyses of archival specimens, in particular with newly available CoV-19 antibody testing, and increased diligence to quantify better CoV-19 testing statistics and test quality measures are needed to evaluate this critical possibility. If these factors are significantly active, they will lead to misinterpretation of current epidemiological data with highly consequential policy missteps.

## Data Availability

All data are publicly available and links to data are referenced in the manuscript.

https://ourworldindata.org/covid-testing

https://www.wwlp.com/news/massachusetts/interactive-map-how-many-coronavirus-tests-have-been-conducted-in-my-state/

